# BurnNet: An Efficient Deep Learning Framework for Accurate Dermal Burn Classification

**DOI:** 10.1101/2021.01.30.21250727

**Authors:** Rohan Bhansali, Rahul Kumar

## Abstract

Burns are the fourth most prevalent unintentional injury around the world, and when left untreated can become permanent and sometimes fatal. An important aspect of treating burn injuries is accurate and efficient diagnosis. Classifying the three primary types of burns – superficial dermal, deep dermal, and full thickness – is essential in determining the necessity of surgery, which is often critical to the afflicted patient’s survival. Unfortunately, reconstructive burn surgeons and dermatologists are merely able to diagnose these types of burns with approximately 50-75% accuracy. As a result, we propose the use of an eight-layer convolutional neural network, BurnNet, for rapid and precise burn classification with 99.87% accuracy. We applied affine transformations to artificially augment our dataset and found that our model attained near perfect metrics across the board, demonstrating the high propensity of deep learning architectures in burn classification.

## 1 Introduction

Burns are a form of human injury caused by the application of high amounts of heat to the skin. Burns are extremely painful as they result in the rapid death of skin cells that can result in permanent damage through the loss of sensation or functionality. They are a prevalent public health issue, ranking as the fourth source of unintentional injury [3].

### 1.1 Dermal Burn Diagnosis

When burns are treated, a determination must be made as to the type and location of the burn for an adequate treatment to occur. The three primary types of burns are superficial dermal burns, deep dermal burns, and full-thickness burns [4]. Superficial burns primarily harm the outer layer of the skin, otherwise known as the epidermis. Typically, they can be described as appearing to be red in color and relatively dry. Blisters may also potentially be present. Deep dermal burns primarily harm the dermis and epidermis layers of the skin and are similar in appearance to superficial burns. Full-thickness burns destroy the dermis, epidermis, and subcutaneous tissue, and generally cause permanent damage. They appear distinctly white and charred. Properly distinguishing between these burns is extremely important in providing pertinent and rapid treatment to burn victims. Unfortunately, in many parts of the world access to specialists is limited and burns are often incorrectly classified, which leads to inadequate treatment measures. In some cases, specialists can take days to be available for consultation which is especially detrimental when surgery is required for treatment. Even when specialists are present, their accuracy can range anywhere from 50% to 76% based on their experience [7].

### 1.2 Previous Deep Learning Research

Convolutional neural networks (CNNs) have been used in the past for burn classification. In one study, CNNs were used to produce feature maps that were fed to a support vector machine (SVM) for classification and an accuracy of 82.43% was achieved [8]. Others have also experimented with CNN architectures for burn degree determination. One research group utilized deep CNNs with local binary operators in order to classify burns as first, second, third, or fourth degree. Notably, it was recorded that the primary features used in burn classifications (by CNNs and surgeons) were color and texture of the burned area [4].

### 1.3 Automated Classification Model

In order to alleviate the problem, we propose BurnNet, a deep convolutional neural network designed to rapidly and accurately classify skin burns. Our research looks to explore the ability of affine transformations to improve the functionality of a deep CNN in burn type classification presented with a small dataset. Specifically, we applied affine transformations in order to artificially magnify our dataset and achieve a training set that is considered adequate in size for deep CNNs. In previous medical imaging work, affine transformations have been shown to have significantly improve results across all metrics, specifically when small datasets were utilized [1]. Additionally, we look to provide clinical viability for burn classification through CNNs by achieving validation accuracy and F1-score above .85, which would be significantly more accurate than most experienced burn surgeons [5].

## 2 Methodology

The dataset that was used in this study was the BIP_US database, containing 94 images of full-thickness, deep dermal, and superficial dermal burns. It was produced by the Signal Theory and Communications Department [2]. The burned area has been cropped and refitted to a gray background.

**Figure 1:**
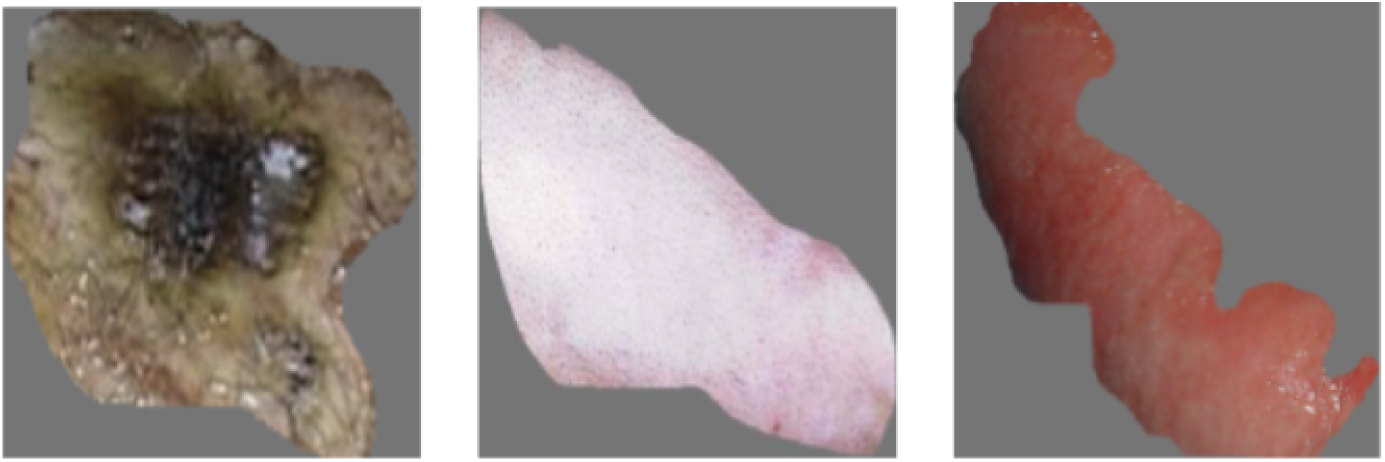
Sample full-thickness, deep dermal, and superficial dermal burn images from BIP_US database

### 2.1 Data Processing

First, all of the images were resized to 512×512 through the anti-aliasing technique. It is among the most sophisticated and robust techniques used in resolution manipulation. It is primarily advantageous because it removes high-frequency noise artifacts prior to resolution manipulation and thus minimizes distortion and black and white noise. Second, we applied affine transformations to all images in the dataset. We applied 72 transformations per image, going in increments of 5 degrees. Examples of the transformed images can be seen below. Next, we split the dataset into a training and testing subset; 75% of images were used for training, with the remainder being used for testing.

### 2.2 BurnNet

The burn classification task is a tertiary classification problem, where the input is an image of a burn and the output is a tertiary label indicating a superficial burn, deep dermal burn, or full-thickness burn, respectively. To accomplish this task, an eight-layer CNN was utilized that we dubbed BurnNet. This model contained an input layer, two convolution layers, two pooling layers, a flattening layer, and two fully connected dense layers. The dense layers alleviate the vanishing gradient problem, strengthen feature propagation, encourage feature reuse, and substantially reduce the number of parameters. Within our model, we utilized Adaptive Moment Estimation (Adam), an adaptive learning rate optimization algorithm, with an initial learning rate of 0.00001. Our activation functions for the intermediate layers was the rectifier function, with the final layer using the softmax function. A batch size of ten was used over twenty epochs, after each of which the model outputted accuracy, performance metrics (precision, recall, and F1 score), and loss, given by the binary cross-entropy loss function:

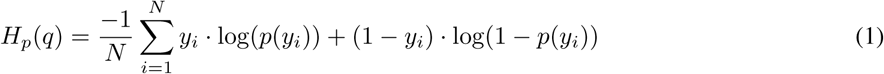

**Figure 2:**
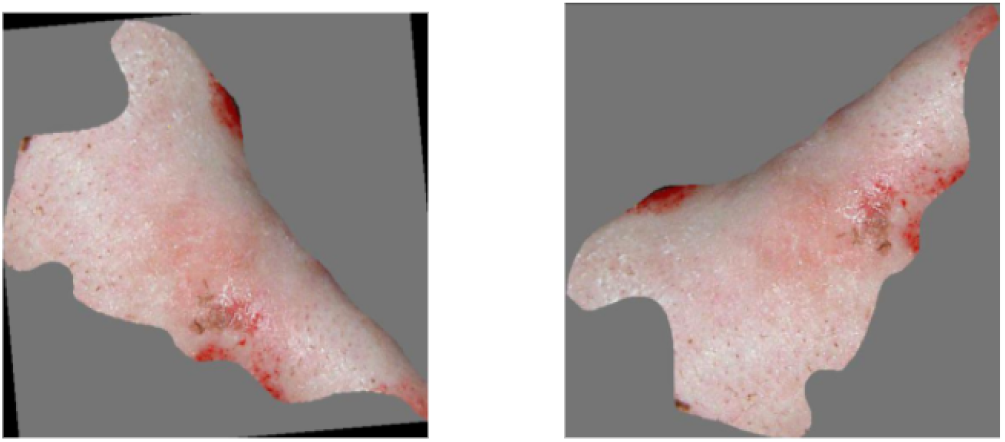
Burn images with affine transformations performed

In order to prevent the networks from overfitting, early stopping was performed by saving the network after every epoch and choosing the saved network with the lowest loss on the tuning set.

**Figure 3:**
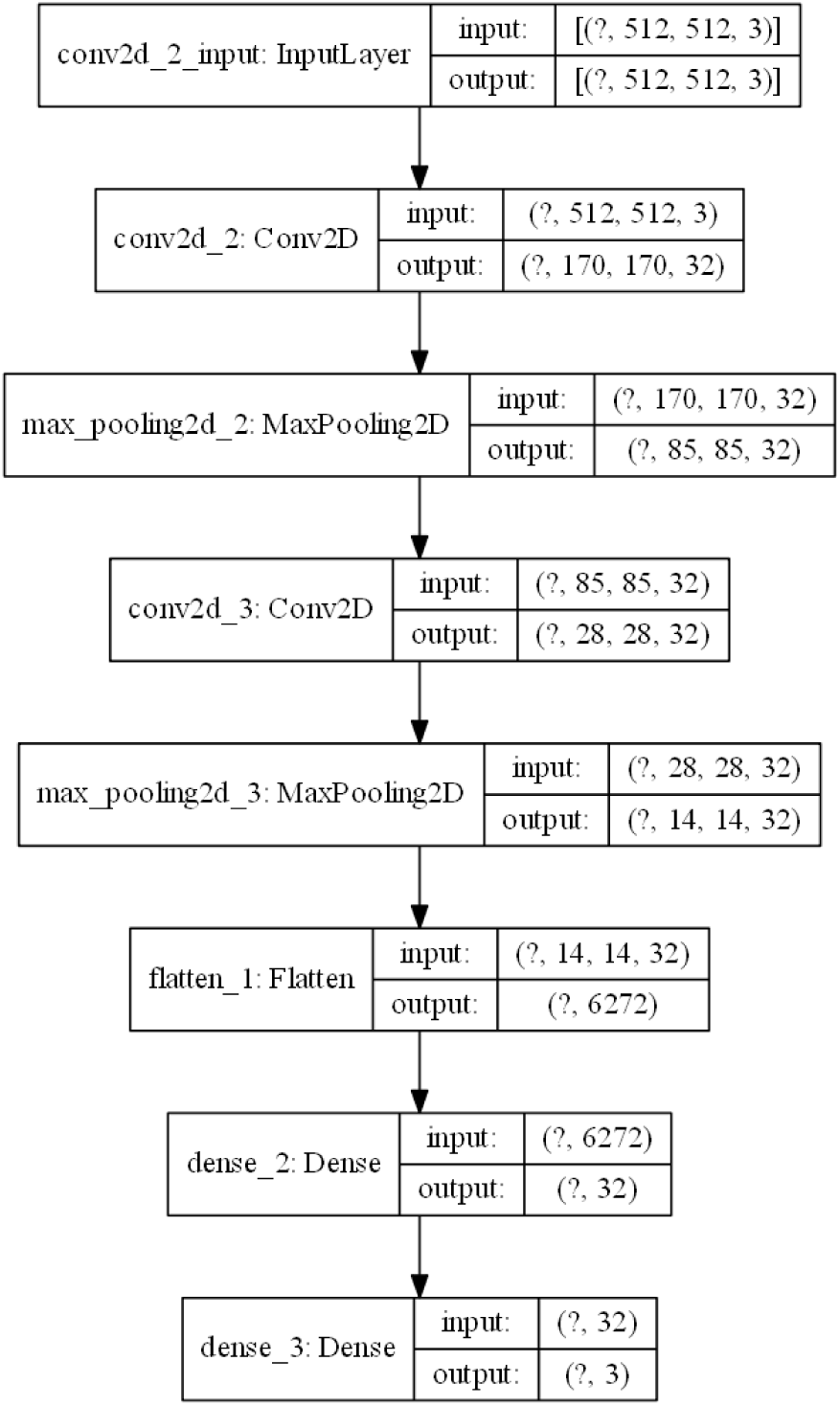
BurnNet Architecture

## 3 Results

Our model attained near-perfect performance metrics across the board, reaching an accuracy, F1, sensitivity, and specificity of 0.9987. We performed early stopping after fifteen epochs to prevent overfitting due to the training loss becoming stagnant.

**Table 1:** Performance Metrics

| Epoch | 5 | 10 | 15 | 20 |
| --- | --- | --- | --- | --- |
| Accuracy | 0.8692 | 0.9590 | 0.9987 | 0.9987 |
| F1 | 0.8667 | 0.9592 | 0.9987 | 0.9987 |
| Sensitivity | 0.8639 | 0.9592 | 0.9987 | 0.9987 |
| Specificity | 0.8698 | 0.9592 | 0.9987 | 0.9987 |

**Figure 4:**
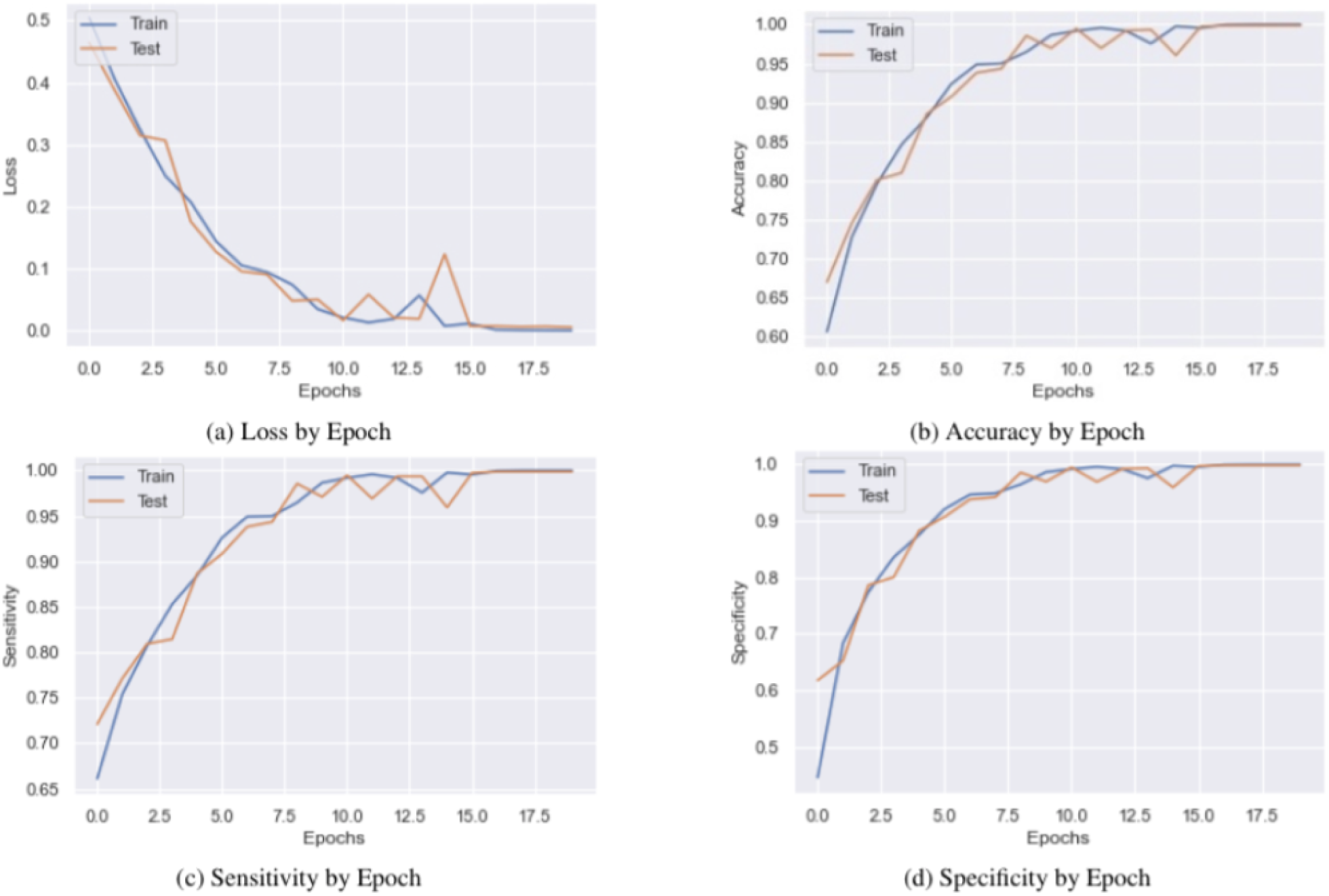
Training Logs

## 4 Conclusion

We utilized affine transformations to artificially augment our image dataset to determine its effect on the performance of our deep convolutional neural network algorithm, BurnNet, to accurately classify images of burned skin as full-thickness, deep dermal, or superficial dermal. Specifically, we applied 72 affine transformations per image in the original dataset and subsequently trained the model for twenty epochs. We found that our model diagnosed dermal burns with 99.87% accuracy, significantly exceeding the efficiency and performance of trained professionals. Our study not only supports the efficacy of deep learning frameworks for dermal burn classification but also provides evidence for the application of data augmentation techniques for improved performance. We hope that our study will provide a major step forward in the automation of medical diagnosis and lower healthcare costs on a global scale.

## Data Availability

The Burn Injury Model System National Database is a prospective, longitudinal, multi-center research data repository that contains measures of functional and psychosocial outcomes following burns. The BMS National Database consists of data collected from individuals with moderate to severe burn injury to learn more about long-term outcomes after a burn injury. BMS data are collected either by paper and pencil, in person or over the phone interviews, or using online surveys. Surveys are completed at discharge from the hospital, 6-months post injury, 12-months post injury, 24-months post injury, and then every five years thereafter. Procedures for data collection are guided by the Standard Operating Procedures. The BMS NDB contains data on over 3,000 adults and almost 2,000 children with moderate to severe burn injury (for a complete list of inclusion criteria, click here). The variables and measures collected include demographics (such as sex, age, race and ethnicity), injury characteristics (such as total body surface area burned and etiology of injury), and outcome measures that assess domains such as pain, itch, depression, and more. Some specific measures currently collected include PROMIS-29, VR-12, and the Post Traumatic Growth Injury.

http://personal.us.es/rboloix/Burns\_BIP\_US\_database.zip

